# Unveiling Chemotherapy’s Impact on Lung Cancer through Single-Cell Transcriptomics

**DOI:** 10.1101/2024.07.09.24310145

**Authors:** Saed Sayad, Mark Hiatt, Hazem Mustafa

## Abstract

**Background:** Lung adenocarcinoma (LUAD) is the most common subtype of non-small cell lung cancer (NSCLC) and frequently affects non-smokers, especially women. It is characterized by a complex genetic profile and interactions with its microenvironment, which contribute to its aggressive and adaptable nature. Early symptoms are often subtle, leading to late diagnoses. Treatment approaches have advanced with targeted therapies and immunotherapy supplementing traditional chemotherapy and radiation. Despite these advancements, the prognosis remains variable, highlighting the need for continued research into new treatment strategies to improve outcomes.

**Method:** In this study, we employed Single-cell RNA Sequencing (scRNA-seq) to comprehensively analyze the impact of chemotherapy on lung adenocarcinoma at the individual cell level. By comparing before and after treatment samples, we assessed the differential expression of genes and pathways, revealing insights into how different cell types within the tumour respond to chemotherapy. This approach enabled us to pinpoint specific mechanisms of drug resistance and highlight potential therapeutic targets for overcoming these challenges.

**Results:** Our analysis uncovered substantial changes in gene expression between primary tumour cells and metastatic cells following chemotherapy. Notably, we observed that 45 pathways were shared between the top 50 upregulated pathways in the primary tumour and the top 50 downregulated pathways in the metastatic tumour post-chemotherapy. Conversely, there was no overlap between the top 50 downregulated pathways in the primary tumour and the top 50 upregulated pathways in the metastatic tumour after chemotherapy. This suggests that chemotherapy effectively downregulated the major upregulated pathways but did not upregulate the key downregulated pathways in metastatic tumours.

**Conclusions:** Integrating single-cell transcriptomics into LUAD research offers detailed insights into the tumour’s response to chemotherapy and its interaction with the immune system. This approach enhances our understanding of LUAD and aids in developing targeted and effective treatments. Based on our analysis, we hypothesize that combining chemotherapy with drugs designed to upregulate the downregulated pathways in primary tumour cells could significantly enhance treatment efficacy and improve patient outcomes.

## Introduction

Lung adenocarcinoma (LUAD) is the most prevalent subtype of non-small cell lung cancer (NSCLC). Unlike other types of lung cancer, adenocarcinoma frequently occurs in non-smokers and is more common in women. The tumour’s complexity is reflected in its diverse genetic landscape and its interaction with the tumour microenvironment, contributing to its aggressive behavior and adaptability. Diagnosis often comes at an advanced stage due to the subtle nature of early symptoms, including cough, chest pain, and shortness of breath. Treatment has evolved significantly, with targeted therapies and immunotherapy complementing traditional approaches like chemotherapy and radiation. Chemotherapy has long stood as a cornerstone in the battle against lung cancer. However, understanding its precise effects on this complex and heterogeneous disease has often been shrouded in mystery [1]. Recent advances in single-cell transcriptomics offer a groundbreaking perspective, providing unprecedented insights into the cellular and molecular changes induced by chemotherapy at an individual cell level. In this exploration, we delve into how single-cell transcriptomics uncovers the dynamic landscape of lung cancer under the influence of chemotherapy.

## Data

We analyzed single-cell RNA sequencing (scRNA-seq) data sourced from the NIH repository (*GSE123902*), as illustrated in Figure 1. Our dataset includes 17 samples collected from 14 distinct patients. All gene-related data has been sourced from the *GeneCards* website, the pathway data from the *Reactome* website and the gene ontology data from the *QuickGO* website.

**Figure 1:**
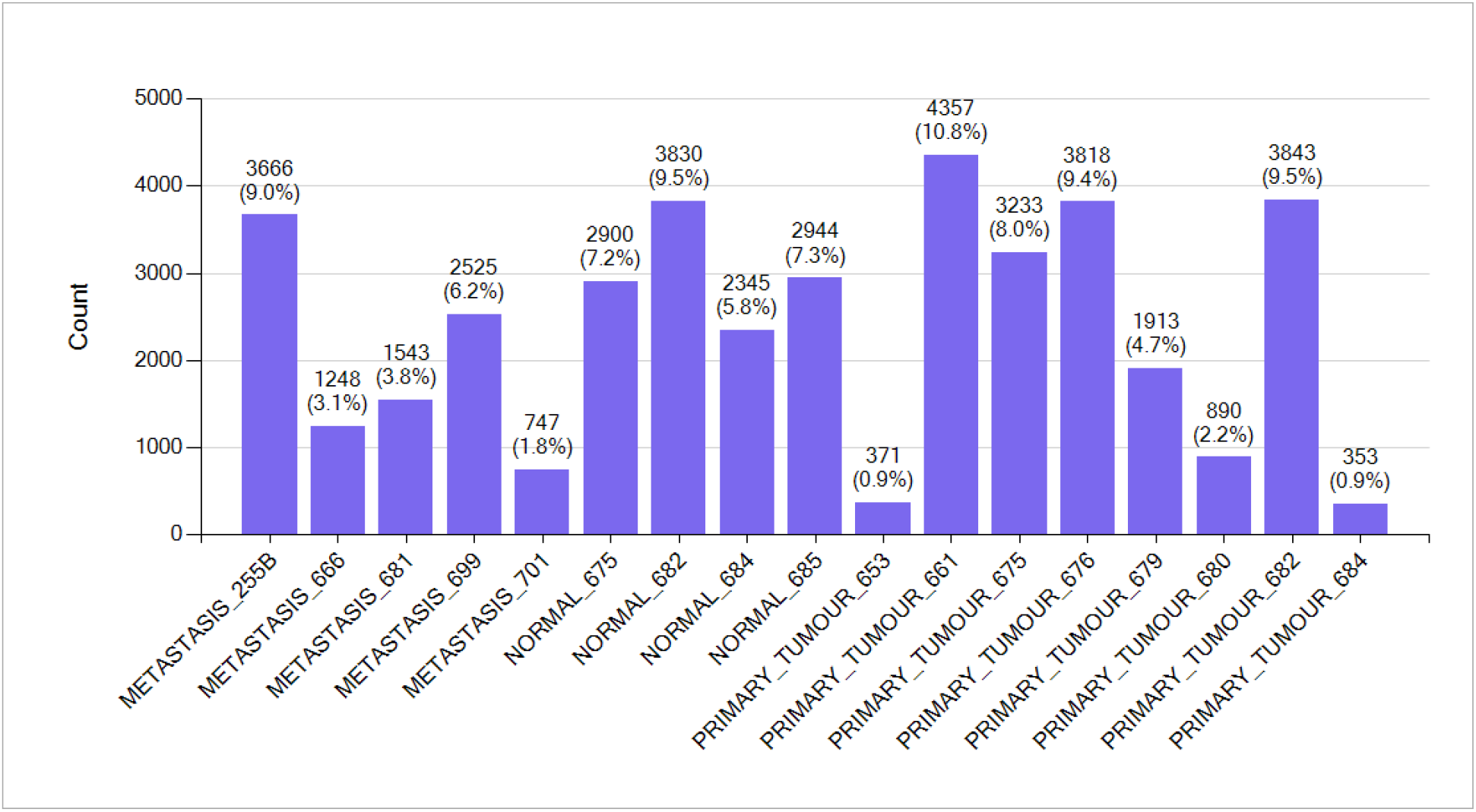
*GSE123902* includes 17 samples collected from 14 distinct patients.

Figure 2 and 3 illustrate the composition of dataset *GSE123902*, encompassing cells from normal lung tissues (NORMAL), primary lung adenocarcinomas (PRIMARY_TUMOUR), and metastases to the brain, bone, and adrenal glands (METASTASIS).

**Figure 2:**
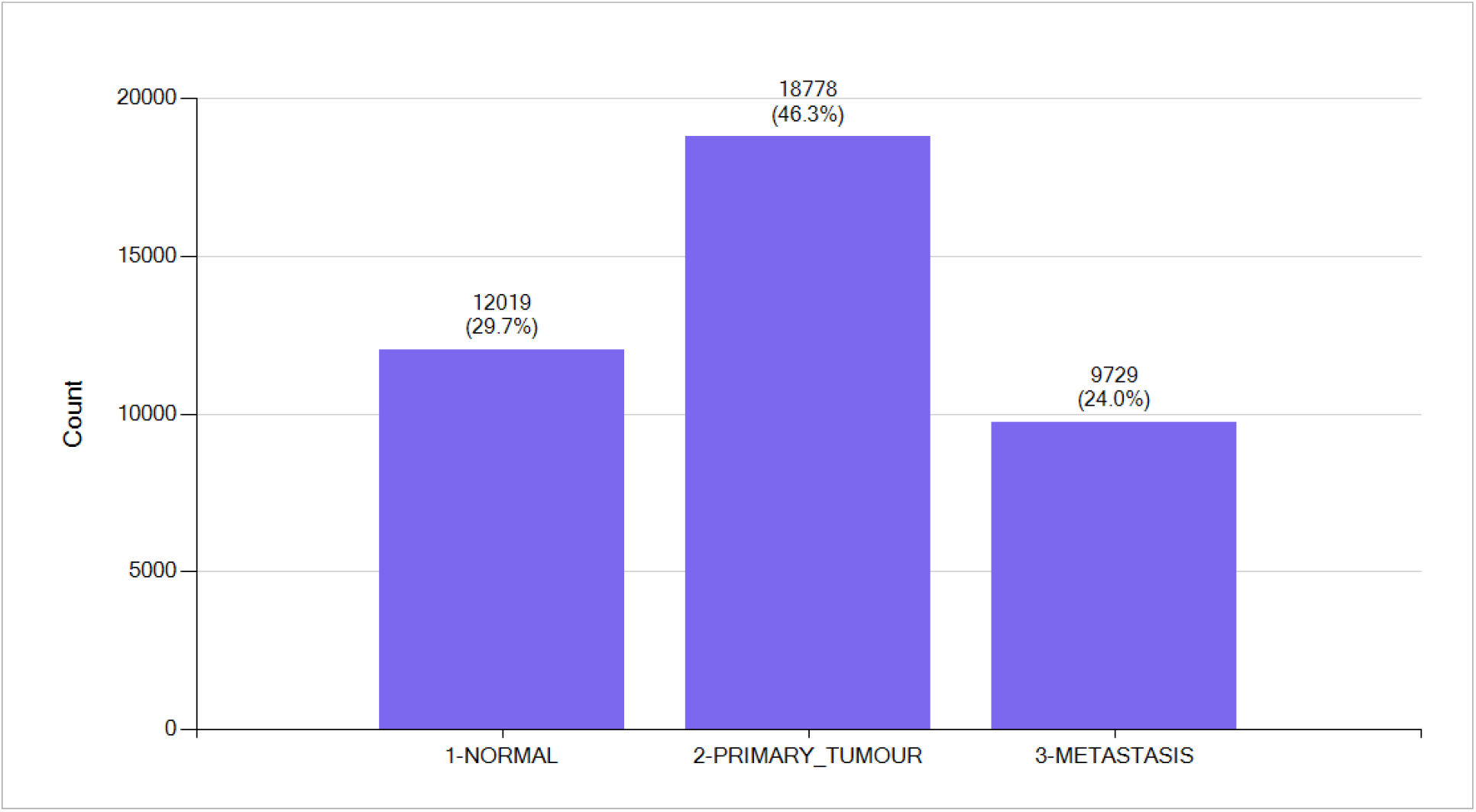
*GSE123902* comprises cells from normal lung tissues (NORMAL), primary lung adenocarcinomas (PRIMARY_TUMOUR), and metastases (METASTASIS) to the brain, bone, and adrenal glands.

**Figure 3:**
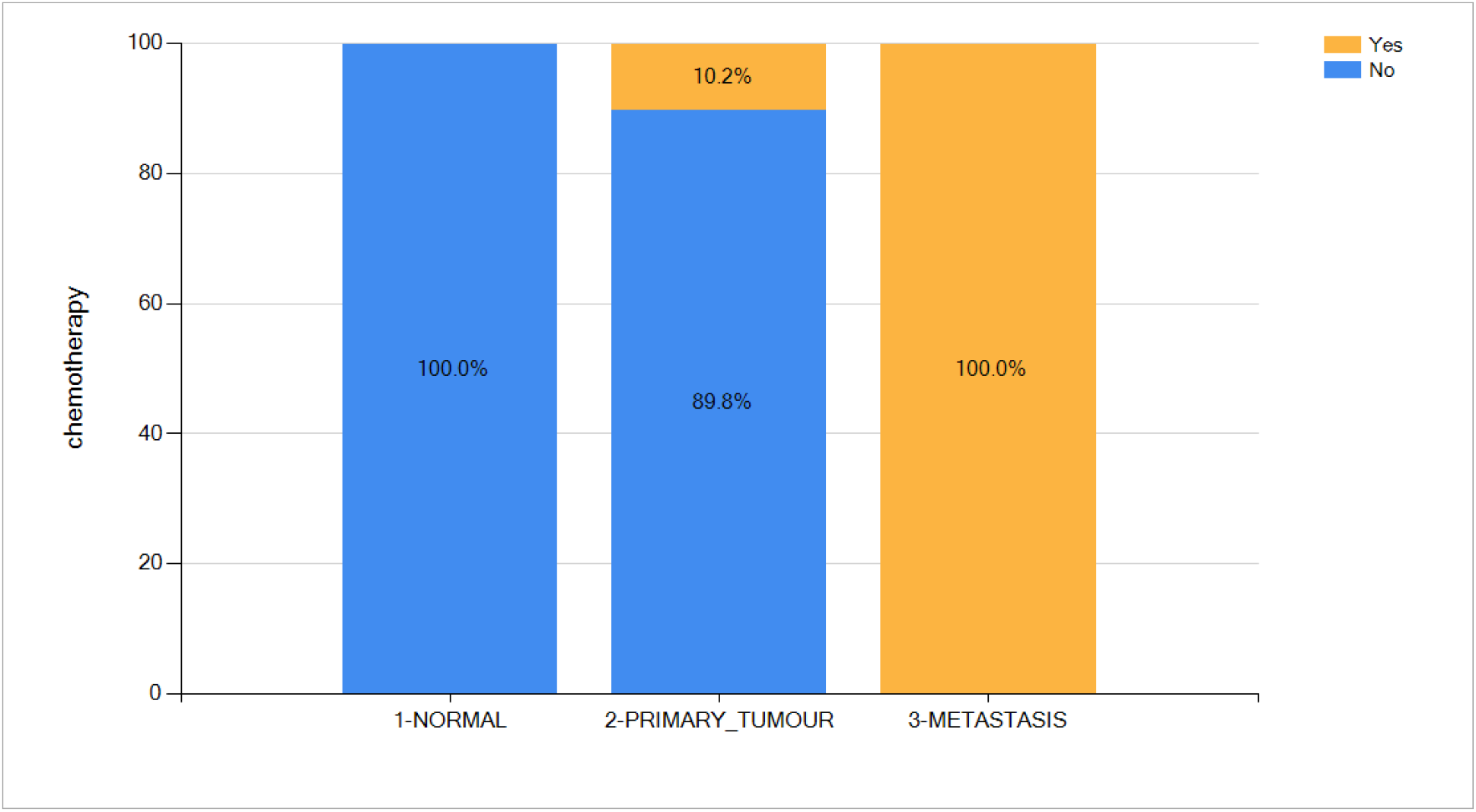
All metastatic samples in the study have undergone chemotherapy treatment.

By profiling scRNA-Seq from 40,526 cells at various stages of lung adenocarcinoma progression, we identified specific genes and pathways that were significantly upregulated or downregulated, especially in response to chemotherapy.

### Differential Gene Expression – NORMAL compared to PRIMARY_TUMOUR

Table 1 presents the top 10 genes that are either downregulated or upregulated when comparing normal to primary tumour single cells. Downregulation and upregulation refer to the decrease or increase in gene expression, respectively.

**Table 1:** Top 10 downregulated and upregulated genes through single-cell gene analysis comparing NORMAL to PRIMARY_TUMOUR samples.

| NORMAL compared to PRIMARY_TUMOUR |  |  |  |
| --- | --- | --- | --- |
| Downregulated in PRIMARY_TUMOUR |  | Upregulated in PRIMARY_TUMOUR |  |
| Gene Symbol | Gene Name | Gene Symbol | Gene Name |
| *FABP4* | Fatty Acid Binding Protein 4 | *IGKC* | Immunoglobulin Kappa Constant |
| *S100A4* | S100 Calcium Binding Protein A4 | *IGHA1* | Immunoglobulin Heavy Constant Alpha 1 |
| *FCGR3A* | Fc Gamma Receptor IIIa | *HSPA1A* | Heat Shock Protein Family A (Hsp70) Member 1A |
| *FGFBP2* | Fibroblast Growth Factor Binding Protein 2 | *IGLC2* | Immunoglobulin Lambda Constant 2 |
| *PRF1* | Perforin 1 | *RGS1* | Regulator Of G Protein Signaling 1 |
| *TYROBP* | Transmembrane Immune Signaling Adaptor TYROBP | *HSPA1B* | Heat Shock Protein Family A (Hsp70) Member 1B |
| *MCEMP1* | Mast Cell Expressed Membrane Protein 1 | *HSP90AA1* | Heat Shock Protein 90 Alpha Family Class A Member 1 |
| *CFD* | Complement Factor D | *DNAJB1* | DnaJ Heat Shock Protein Family (Hsp40) Member B1 |
| *NKG7* | Natural Killer Cell Granule Protein 7 | *HSPE1* | Heat Shock Protein Family E (Hsp10) Member 1 |
| *GLIPR2* | GLI Pathogenesis Related 2 | *HSPH1* | Heat Shock Protein Family H (Hsp110) Member 1 |

The top 10 downregulated genes exhibit reduced expression in primary tumours, with significant implications for immune response, cell signaling, and cellular defense mechanisms. Among these, *FABP4* [2], *S100A4* [3], and *FCGR3A* [4] are notably associated with these crucial biological processes. Key downregulated genes such as *PRF1* [5] and *TYROBP* [6] play vital roles in the cytolytic activity of immune cells and their activation, respectively. In lung cancer, the fibroblast growth factor receptor (FGFR) pathway is pivotal in signal transduction. It regulates a wide range of cellular processes, including cell cycle progression, migration, metabolism, survival, proliferation, and differentiation [7]. Additionally, *MCEMP1* deficiency has been shown to impair stem cell factor-induced peritoneal mast cell proliferation in vitro and reduce lung mast cell expansion in vivo [8]. GLI pathogenesis-related 1 (GLIPR1) acts as a tumour suppressor in lung cancer, underscoring its potential as a therapeutic target [9]. The role of the complement system in lung cancer progression is also highlighted by its signaling interactions with immune cells within the tumour microenvironment [10]. Furthermore, natural killer (NK) cells in the lung cancer microenvironment demonstrate reduced cytotoxicity, impaired viability, and a distinct phenotype, indicating a shift away from their typical immune functions and suggesting potential immune evasion strategies or altered metabolic needs within the primary tumour environment [11].

Conversely, the top 10 upregulated genes exhibit increased expression in primary tumours and are predominantly associated with the immune system and cellular stress responses. For instance, *IGKC, IGHA1*, and *IGLC2* are parts of immunoglobulins essential for immune defense and emerging evidence suggested that the production of immunoglobulin has been detected in various cancer cells, such as lung adenocarcinoma and macrophages. Cancer-derived immunoglobulin has been implicated in promoting tumour proliferation and facilitating immune escape [12]. Additionally, *HSPA1A, HSPA1B, HSP90AA1, HSPE1, HSPH1* and *DNAJB1* are heat shock proteins that help cells cope with stress by assisting in protein folding and stabilization and have been reported to be associated with the survival time of lung cancer patients, which may be a potential biomarker in lung cancer treatment [13]. Furthermore, *RGS1* modulates autophagic and metabolic programs and is a critical mediator of human regulatory T cell function [14].

### Differential Gene Expression – PRIMARY_TUMOUR compared to METASTASIS + chemotherapy

Table 2 highlights the top 10 genes that are either downregulated or upregulated when comparing primary single cells to metastatic single cells after chemotherapy treatment. A notable finding is that three heat shock genes—*HSPA1A, HSP90AA1*, and *HSPH1*—exhibit a reversal in their expression patterns: they are upregulated in the primary tumour but significantly downregulated in the metastatic tumour following chemotherapy. Furthermore, our analysis reveals 33 genes that are common between the top 100 upregulated genes in the primary tumour and the top 100 downregulated genes in the metastatic tumour post-chemotherapy. Conversely, no overlap is observed among the top 10 downregulated genes in the primary tumour and the upregulated genes in the metastatic tumour post-chemotherapy. Among the top 100 downregulated genes in the primary tumour, only two are found to be upregulated in the metastatic tumour post-treatment. It indicates that chemotherapy affects gene expression patterns in a selective manner. This variability suggests that not all genes associated with tumour growth or metastasis respond uniformly to chemotherapy, which may influence treatment efficacy and patient outcomes.

**Table 2:** Top 10 downregulated and upregulated genes through single-cell gene analysis comparing PRIMARY_TUMOUR to METASTASIS samples following chemotherapy.

| PRIMARY_TUMOUR compared to METASTASIS + chemotherapy |  |  |  |
| --- | --- | --- | --- |
| Downregulated in METASTASIS + chemotherapy |  | Upregulated in METASTASIS + chemotherapy |  |
| Gene Symbol | Gene Name | Gene Symbol | Gene Name |
| *BTG1* | BTG Anti-Proliferation Factor 1 | *CMBL* | Carboxymethylenebutenolidase Homolog |
| *CXCR4* | C-X-C Motif Chemokine Receptor 4 | *SOD3* | Superoxide Dismutase 3 |
| *CD69* | CD69 Molecule | *MDK* | Midkine |
| *HSPA1A* | Heat Shock Protein Family A (Hsp70) Member 1A | *FASN* | Fatty Acid Synthase |
| *HSP90AA1* | Heat Shock Protein 90 Alpha Family Class A Member 1 | *MUC4* | Mucin 4, Cell Surface Associated |
| *HSPH1* | Heat Shock Protein Family H (Hsp110) Member 1 | *SAA1* | Serum Amyloid A1 |
| *TXNIP* | Thioredoxin Interacting Protein | *SFTPB* | Surfactant Protein B |
| *IL7R* | Interleukin 7 Receptor | *CKB* | Creatine Kinase B |
| *HSPA8* | Heat Shock Protein Family A (Hsp70) Member 8 | *MIR4458HG* | MIR4458 Host Gene |
| *KLF6* | KLF Transcription Factor 6 | *CLPTM1L* | CLPTM1 Like |

**Table 3:** Top 10 downregulated and upregulated pathways identified through single-cell gene enrichment analysis comparing NORMAL to PRIMARY_TUMOUR samples.

| NORMAL compared to PRIMARY_TUMOUR |  |
| --- | --- |
| Downregulated in PRIMARY_TUMOUR | Upregulated in PRIMARY_TUMOUR |
| FGFR2b ligand binding and activation | vRNP Assembly |
| FGFR2 ligand binding and activation | Resistance of ERBB2 KD mutants to trastuzumab |
| Triglyceride catabolism | Resistance of ERBB2 KD mutants to afatinib |
| Triglyceride metabolism | Resistance of ERBB2 KD mutants to tesevatinib |
| Alternative complement activation | Resistance of ERBB2 KD mutants to neratinib |
| Pregnenolone biosynthesis | Drug resistance in ERBB2 KD mutants |
| Metabolism of serotonin | Drug resistance in ERBB2 TMD/JMD mutants |
| Serotonin clearance from the synaptic cleft | Resistance of ERBB2 KD mutants to AEE788 |
| Neurotransmitter clearance | Resistance of ERBB2 KD mutants to sapitinib |
| Ethanol oxidation | Resistance of ERBB2 KD mutants to lapatinib |

### Pathways Analysis – NORMAL compared to PRIMARY_TUMOUR

In the context of comparing normal single cells to primary tumour single cells, the top downregulated pathways in primary tumours primarily involve metabolic and neurotransmitter processes. For instance, the *fibroblast growth factor receptor* (FGFR) pathway plays a key role in signal transduction in lung cancer. It controls cellular processes such as cell cycle progression, migration, metabolism, survival, proliferation, and differentiation [15]. Moreover, pathways such as *Triglyceride catabolism, Triglyceride metabolism*, and *Ethanol oxidation* are less active in tumour tissues, indicating a potential shift in energy utilization or metabolic processes within the tumour environment. Moreover, fatty acid metabolism plays a significant role in tumourigenesis, progression, and immune regulation [16]. Additionally, pathways related to ueurotransmitters, such as *Metabolism of serotonin, Serotonin clearance from the synaptic cleft and Neurotransmitter clearance*, suggesting alterations in neurotransmitter management that could impact cellular signaling and modulate the tumour microenvironment [17]. Furthermore, downregulation of *Pregnenolone biosynthesis* demonstrates changes in steroid hormone synthesis and immune response in primary tumours [18].

On the other hand, the top upregulated pathway, *vRNP Assembly*, which is related to the assembly of viral ribonucleoprotein complexes, potentially reflecting an increased need for viral-like processes or mimicry in tumour cells. Viruses have been central to modern cancer research and provide profound insights into cancer causes but the role of virus in lung cancer is still unclear [19]. Additionally, the majority of upregulated pathways in primary tumours involve mechanisms related to drug resistance and signaling via the ERBB2 (HER2) receptor. These include various pathways linked to *Resistance of ERBB2 KD mutants* to several inhibitors like trastuzumab, afatinib, and neratinib, indicating a robust mechanism of resistance in cancer cells. ERBB2 amplification events in non-small cell lung cancer (NSCLC) are observed in two distinct clinicopathologic and genomic contexts. In patients with a smoking history, ERBB2 often acts as the primary mitogenic driver. However, in patients with a light or never smoking history, ERBB2 amplification frequently co-occurs with other mitogenic drivers [20].

### Pathways Analysis – PRIMARY_TUMOUR compared to METASTASIS + chemotherapy

Table 4 showcases the top 10 pathways that exhibit significant changes in expression when comparing primary single cells to metastatic single cells post-chemotherapy. Notably, 8 out of these 10 pathways demonstrate a striking reversal in their expression patterns: they are upregulated in the primary tumour but become significantly downregulated in the metastatic tumour after chemotherapy treatment. Our analysis also identifies 45 pathways that overlap between the top 50 upregulated pathways in the primary tumour and the top 50 downregulated pathways in the metastatic tumour following chemotherapy. In contrast, there is no overlap among the top 50 downregulated pathways in the primary tumour and the top 50 upregulated pathways in the metastatic tumour post-chemotherapy. This variability highlights that pathways linked to tumour growth and metastasis do not uniformly respond to chemotherapy, which may have significant implications for treatment effectiveness and patient outcomes.

**Table 4:**
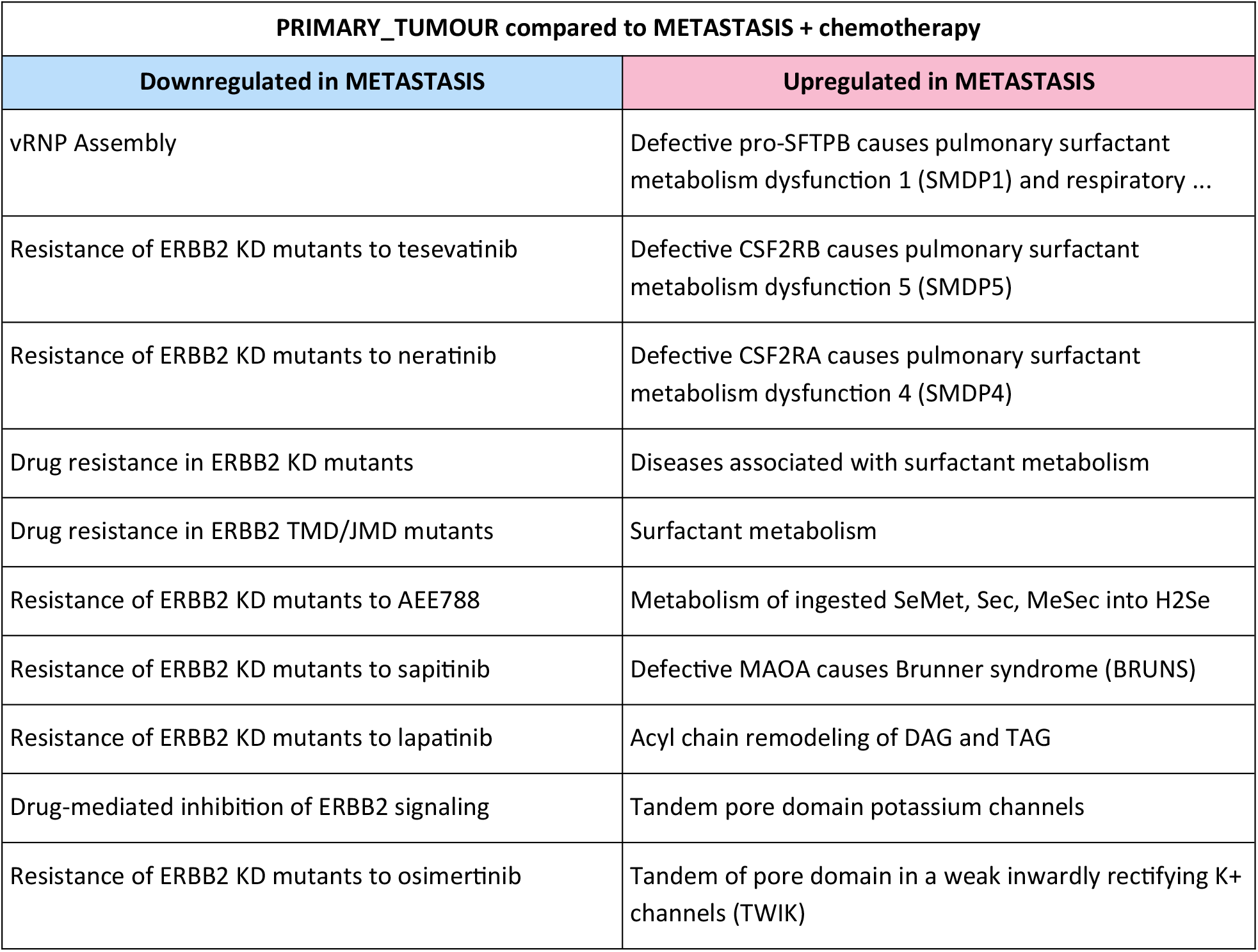
Top 10 pathways showing downregulation and upregulation identified through single-cell gene enrichment analysis comparing PRIMARY_TUMOUR to METASTASIS samples following chemotherapy.

## Discussion

The application of scRNA-seq in studying lung adenocarcinoma (LUAD) has unveiled critical insights into the cellular and molecular underpinnings of this aggressive cancer, especially in response to chemotherapy. This study harnesses the power of scRNA-seq to dissect the complex genetic landscape of LUAD at a resolution previously unattainable, allowing us to observe how individual cells adapt to and resist therapeutic interventions. One of the key revelations of our analysis is the heterogeneous response of LUAD cells to chemotherapy, a cornerstone in lung cancer treatment. The selective changes in gene expression, particularly the reversal patterns observed in heat shock proteins such as HSPA1A and HSPH1 between primary and metastatic tumours post-chemotherapy, underscore the adaptive mechanisms that cancer cells employ to survive. This adaptive response is not uniform across all genes, indicating that chemotherapy induces a spectrum of cellular responses rather than a monolithic shift in gene expression. Such findings highlight the need for personalized therapeutic approaches that can target these diverse responses more effectively.

The differential gene expression analysis between normal and primary tumour cells highlights significant dysregulation in genes associated with immune response and cellular stress. For example, the upregulation of heat shock proteins and immunoglobulins in primary tumours points to the critical role of these proteins in supporting tumour cell survival and possibly aiding in immune evasion. Conversely, the downregulation of genes involved in immune cell activation and cytolytic activity, such as PRF1 and TYROBP, suggests a compromised immune surveillance in the tumour microenvironment. This dual perspective of upregulated survival mechanisms and downregulated immune defense provides a comprehensive view of how LUAD manipulates its environment to promote growth and resist immune attacks.

Pathway analysis further complements these findings by illustrating the metabolic and signaling shifts between normal and cancerous states. The downregulation of metabolic pathways and the upregulation of pathways related to drug resistance and viral-like processes in primary tumours align with the metabolic reprogramming often seen in cancer cells. These insights into altered metabolic processes provide potential targets for disrupting the energy and resource utilization of cancer cells, possibly enhancing the efficacy of existing therapies.

Moreover, the differential pathway expression between primary tumours and metastatic sites following chemotherapy treatment sheds light on the dynamic and adaptive nature of cancer progression. The significant overlap in pathways that are upregulated in primary tumours and downregulated in metastatic tumours after chemotherapy suggests that certain cellular functions are critical during early tumour growth but become less advantageous or even detrimental as the tumour metastasizes and faces therapeutic pressure. This adaptability highlights the challenge of treating metastatic cancer, where the molecular landscape can shift significantly, rendering some treatments less effective. We hypothesize that combining chemotherapy with drugs designed to upregulate the downregulated genes in primary tumour cells could significantly enhance treatment efficacy and improve patient outcomes.

## Summary

Integrating single-cell transcriptomics into lung adenocarcinoma (LUAD) research has provided detailed insights into the tumour’s response to chemotherapy and its interaction with the immune system, enhancing our understanding of LUAD and aiding in the development of more targeted treatments. Our study identified significant gene expression changes between primary and metastatic tumour cells following chemotherapy. We discovered that 45 pathways overlapped between the top 50 upregulated pathways in primary tumours and the top 50 downregulated pathways in metastatic tumours, indicating that chemotherapy downregulated the key pathways in primary tumours. However, there was no overlap in the opposite direction, suggesting that chemotherapy did not upregulate the major downregulated pathways in metastatic tumours. Based on these findings, we hypothesize that combining chemotherapy with drugs designed to activate the downregulated pathways in primary tumours could significantly improve treatment efficacy and patient outcomes.

## Data Availability

All data produced are available online at:
https://www.ncbi.nlm.nih.gov/geo/query/acc.cgi?acc=GSE123902

